# Household Secondary Attack Rate in Gandhinagar district of Gujarat state from Western India

**DOI:** 10.1101/2020.09.03.20187336

**Authors:** Komal Shah, Nupur Desai, Deepak Saxena, Dileep Mavalankar, Umang Mishra, G C Patel

**Affiliations:** Indian Institute of Public Health Gandhinagar - 382042, Gujarat, India; New York University, USA; Epidemic, Commissioner (Health, Medical Services and Medical Education), Gandhinagar; Epidemic Branch, Commissionerate of Health, Gandhinagar, Gujarat

## Abstract

Objectives: Current retrospective study aims to evaluate household Secondary Attack Rate (SAR) of COVID-19 in Gandhinagar (rural) district of Gujarat, India. Methods: Line-listing of 486 laboratory-confirmed patients, tested between 28th March to 2nd July was collected, out of them 80 (15% of overall sample) cases were randomly selected. Demographic, clinical and household details of cases were collected through telephonic interview. During interview 28 more patients were identified from the same household and were added accordingly. So, study included 74 unrelated cluster of households with 74 primary cases and 386 close contacts. Results: SAR in household contacts of COVID-19 in Gandhinagar was 8.8%. Out of 108, 8 patients expired (7.4%), where higher mortality was observed in primary cases (9.5%) as compared to secondary cases (3%). Occupational analysis showed that majority of the secondary cases (88%) were not working and hence had higher contact time with patient. No out-of-pocket expenditure occurred in 94% of the patients, in remaining 6% average expenditure of 1,49,633INR (2027 USD) was recorded. Conclusions: Key observations from the study are 1) SAR of 8.8% is relatively low and hence home isolation of the cases can be continued 2) Primary case is more susceptible to fatal outcome as compared to secondary cases 3) Government has covered huge population of the COVID-19 patients under cost protection. However, more robust studies with larger datasets are needed to further validate the findings.

## 1. Introduction

On global chart of COVID-19, India stands at 3^rd^ position with 2.4 million cases and around 47,000 deaths as per the latest reports published on 13^th^ August, 2020^1^. By far transmission trends showed that containment, contact tracing and surveillance are the effective strategies for limiting the spread of infection^2,3^. Studies have showed that transmission probabilities are highest among household contacts with greater vulnerability of spouses and elderly^4,5^. It was also observed that close contacts having comorbid conditions are at higher risk of secondary infection^6,7^. Recently conducted two systematic reviews studied characteristic features of COVID-19 transmission in household contacts^8,9^. Both showed that household secondary attack rate (SAR) varies widely among different populations and ranges from 4.6% to 49%.

India is a culturally, genetically, environmentally and geographically diverse country with specific disease determinants. Hence population specific understanding of the disease transmission is vital to design country specific guidelines. To the best of our knowledge till date there are only two studies published from India that specifically studied SAR^10,11^. A national study published by Indian Council of Medical Research showed SAR of 6% (national average) with highest rate in Chandigarh (11.5%) and Maharastra (10.6%) state. Apart from this Ramanan et al published SAR from two Indian states – Tamil Nadu and Andhra Pradesh and reported it to be 9% (7.5% - 10.5%). The authors showed that as compared to community contact the household contact have 3.56 (2.99 – 4.22) times higher risk of secondary transmission, even after adjusting other risk factors. Both the studies have emphasized the need of robust primary data to design locally-appropriate control measures.

With current retrospective study, we aim to assess household SAR in Gandhinagar district of Gujarat state from Western India with an intend to study prevalence, determinants and cost associated with household Secondary attack. Gandhinagar is the capital district of State of Gujarat with city population of 4,87,392 and 216 villages with 9,04,361 of population.

## 2. Methods

For the study data set of laboratory confirmed COVID-19 patients from Gandhinagar, rural district was obtained from Government records. The line list consisted of 486 cases who were diagnosed positive between 28^th^ March to 2^nd^ July, 2020. For the study 15% of the positive cases (n = 80) were randomly selected, through the computerized method of random sequence generation from the provided list. Those 80 cases were from unrelated cluster of 74 households. An interview tool was developed to collect the information regarding demographic, clinical and household details from the cases through telephonic interview. The study was reviewed and approved by Gujarat Government and Institutional Ethics Committee (IEC). Detailed interview of the selected 80 cases resulted into identification of 28 new cases who were either primary or secondary cases of the initially selected patients. So, the overall study included 108 cases from 74 households where 74 were primary and 34 were secondary cases of COVID-19 from 386 close household contacts.

### Data collection

Demographic, clinical, household, comorbid conditions and cost of diagnosis and treatment of primary and secondary cases were collected using pre-validated data collection tool (figure 1). Initially the tool was validated in few cases and was modified based upon experience of this validation exercise. The patients were approached for the study through telephonic interview. After obtaining verbal consent the details were collected from each primary and secondary case. Household contact was defined as contact sharing same residential address.

**Figure 1:**
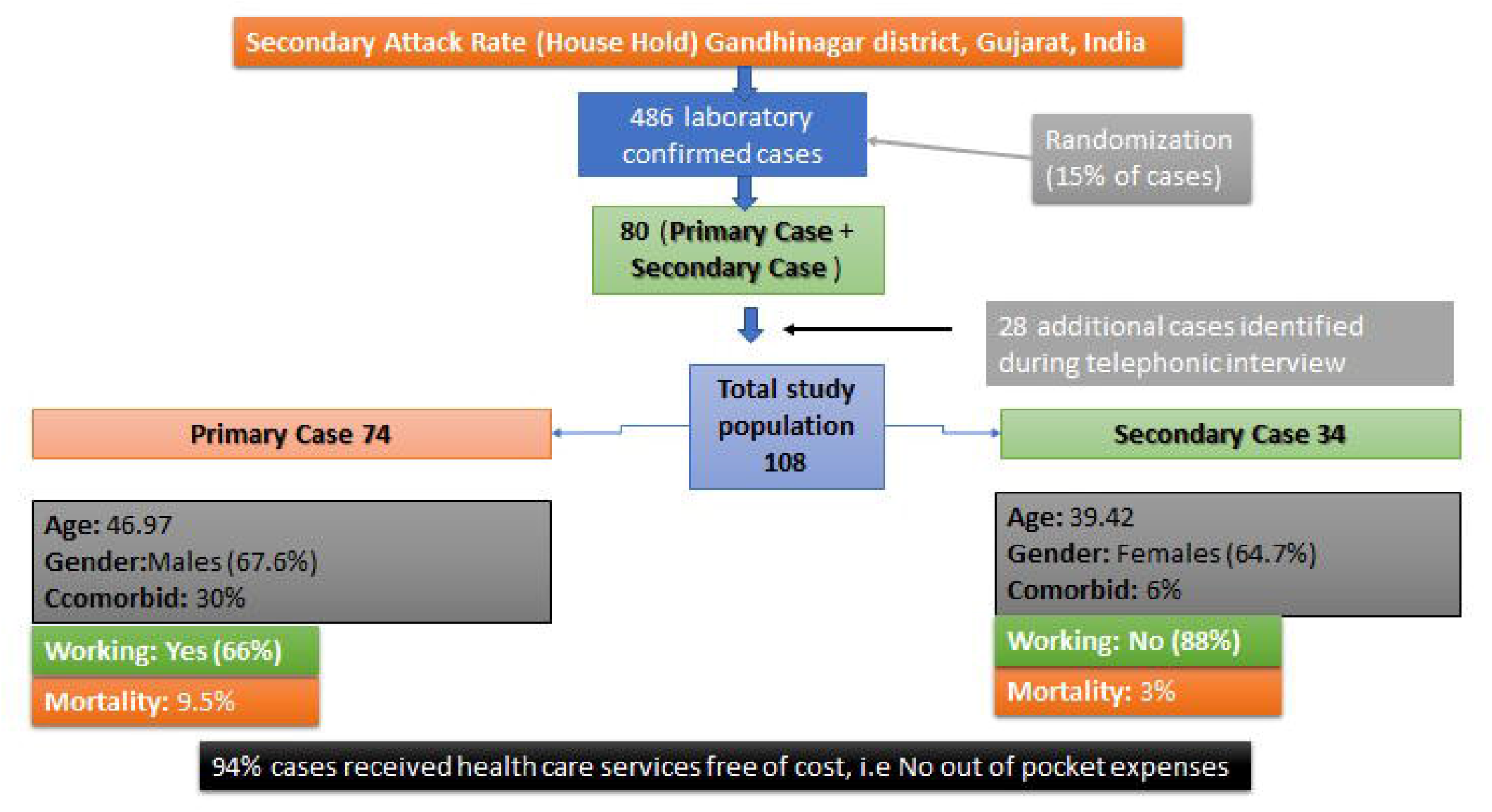
Diagrammatical representation of the study methodology and outcome

## 3. Results

Key findings of the study regarding deaths in primary and secondary cases and household SAR are presented in table 1. The diagrammatic representation of the entire study protocol and important observation are presented in figure 1. One of the significant finding was that we observed 0% drop out rate in the study. Household SAR was calculated as a number of household cases occurring within the 28 days incubation time after exposure to a primary case divided by total susceptible household contacts. Out of 386 household contacts of 74 primary cases, 34 contacts developed secondary infection and hence SAR was 8.8% in the studied population. All the enrolled primary and secondary cases were hospitalized and were confirmed through RT-PCR test. Overall death experienced in primary cases were higher as compared to secondary cases (9.5% vs 3%; p = 0.23).

**Table 1:**
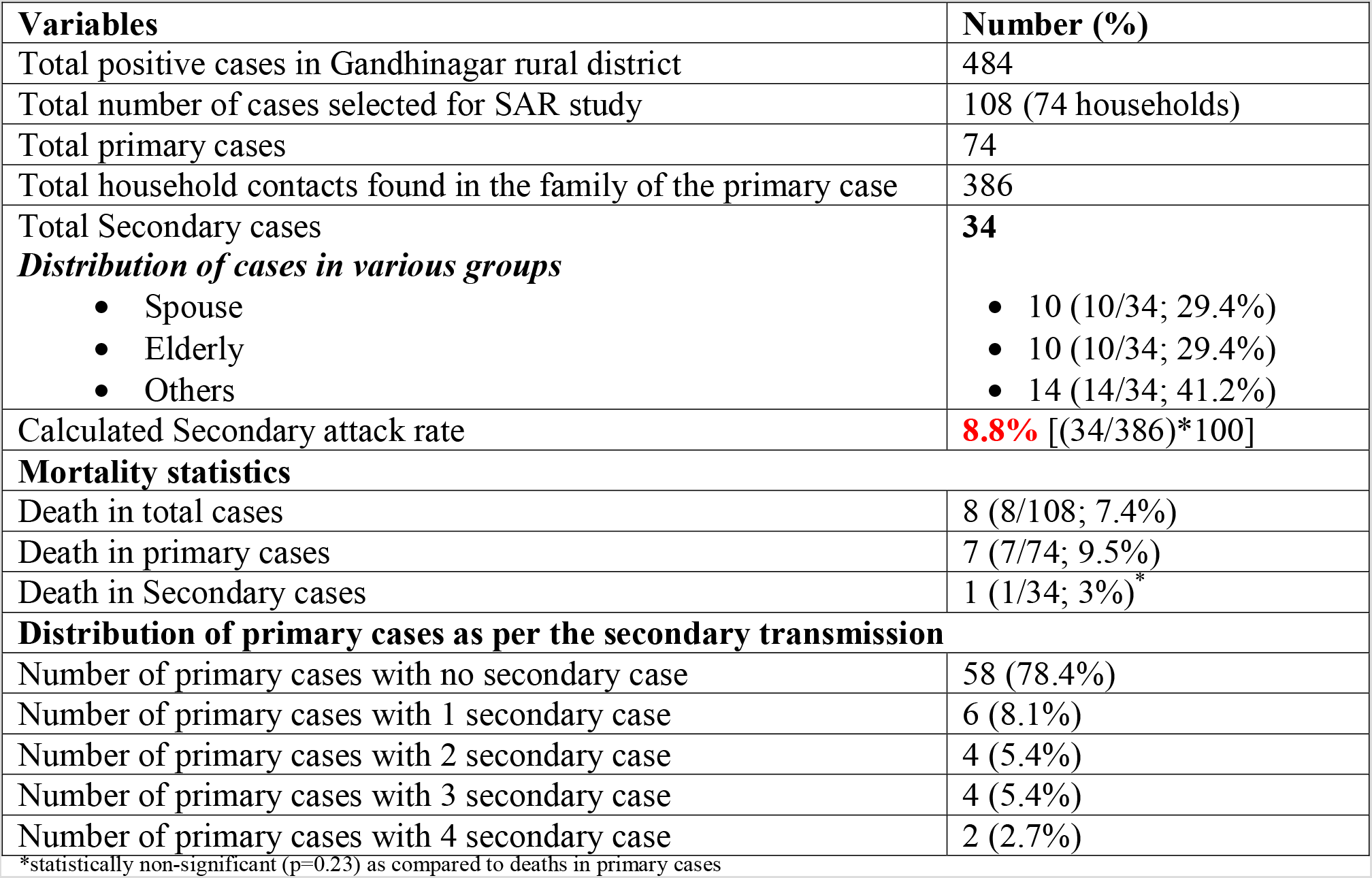
Details of study population, SAR and mortality in Gandhinagar district

| **Variables** | **Number (%)** |
| --- | --- |
| Total positive cases in Gandhinagar rural district | 484 |
| Total number of cases selected for SAR study | 108 (74 households) |
| Total primary cases | 74 |
| Total household contacts found in the family of the primary case | 386 |
| Total Secondary cases | **34** |
| ***Distribution of cases in various groups*** |  |
| • Spouse | • 10 (10/34; 29.4%) |
| • Elderly | • 10 (10/34; 29.4%) |
| • Others | • 14 (14/34; 41.2%) |
| Calculated Secondary attack rate | **8.8%** [(34/386)*100] |
| **Mortality statistics** |  |
| Death in total cases | 8 (8/108; 7.4%) |
| Death in primary cases | 7 (7/74; 9.5%) |
| Death in Secondary cases | 1 (1/34; 3%)* |
| **Distribution of primary cases as per the secondary transmission** |  |
| Number of primary cases with no secondary case | 58 (78.4%) |
| Number of primary cases with 1 secondary case | 6 (8.1%) |
| Number of primary cases with 2 secondary case | 4 (5.4%) |
| Number of primary cases with 3 secondary case | 4 (5.4%) |
| Number of primary cases with 4 secondary case | 2 (2.7%) |

\*statistically non-significant (p=0.23) as compared to deaths in primary cases

As shown in table 2 mean age of primary cases was higher than secondary cases. The secondary cases were predominated by female patients (65%). However, prevalence of comorbid conditions was low in secondary cases as compared to primary cases. Occupational analysis showed that 67.6% of the primary cases were working outside their home and hence possibly caught the infection from sources outside home. More females were infected from the primary cases(64.7%) and majority of the secondary cases were not having any occupation (88.2%) and were involved in household work only. This indicates that the potential source of infection transmission was primary case and not any other source. It also indicates that the contacts developing secondary infection might be spending more time with the primary contact. Primary cases were further grouped as per the infected secondary cases (table 2). It was found that only 6 primary cases (8.1%) infected 3 or more secondary cases in the household contacts.

**Table 2:**
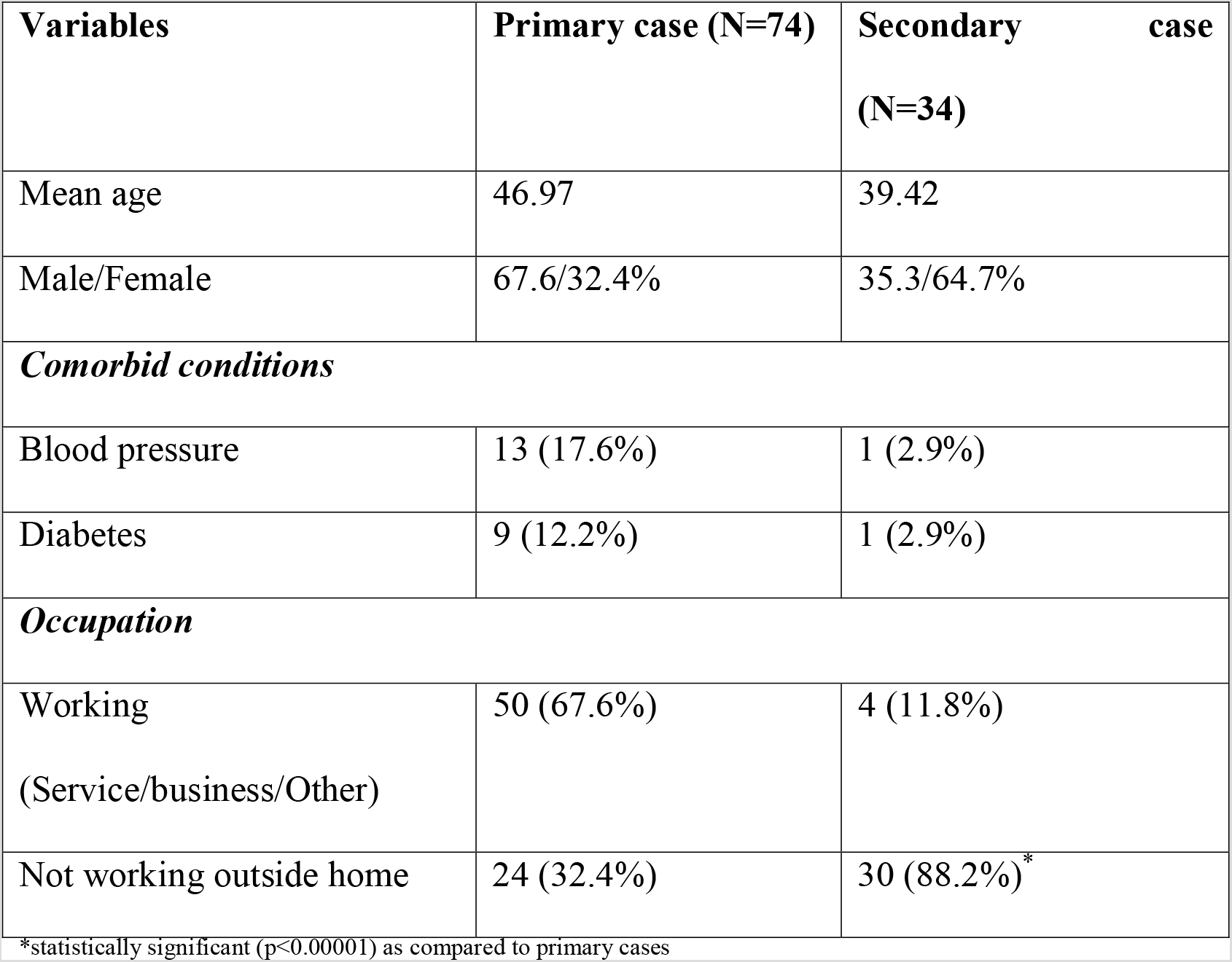
Demographic and clinical details of primary and secondary cases in Gandhinagar district

Majority of the patients (94%) received health care facilities including diagnosis and treatment free of cost in government facilities, however few of them (6%) used private health care facilities and the average cost was 1,49,633 INR (2027 USD) in them. Thus, government has provided great subsidy/ cost protection by providing completely free services.

## 4. Discussion

It is reported by various studies that household contacts of COVID-19 patients are at greater risk of developing disease as compared to other contacts, principally due to higher contact time in home^12,13^. Also, during household contact general preventive measures such as wearing of mask and physical distancing norms are often not followed.

In different states of India COVID-19 statistics are relatively different than the other states and so the SAR prevalence and its factors were also expected to be different. Hence estimating SAR in various districts and cities or even wards of city is very important.

Current study provided some important insights into transmission of the disease in household contacts in Gandhinagar district of Gujarat State. When compared with ICMR national statistics where Gujarat had SAR of 7.8%, our study showed relatively similar SAR from rural setting (8.8%), however this needs to be interpreted with caution as current study included data from only one districts of Gujarat and may not be a representative data of entire state. One recent global review conducted by same team of researchers summarized that SAR varies widely across countries with lowest reported rate as 4.6% and highest as 49.56%. In the same line ICMR study representing Indian statistics also showed range of 0–11.5%, with a national average of 6%^10^.

Though the study was conducted in only one districts and the more data are needed for generalizability of the findings, it was indicated that mortality in primary cases are comparatively higher than secondary cases. However, it did not reach to a statistically significant level and greater sample size with higher event rate is needed to substantiate this further.

Categorization of the case according to occupation showed two key findings: 1) In primary cases the individuals working in service sector constituted more than half of the group indicating possibility of catching the infection outside house 2) Secondary cases were majorly found in contact who are not engaged in any work outside houses and hence are expected to spend greater time with the primary contact. It also indicated that there are higher chances of them getting transmission of virus through primary case only.

One important find of the study is that availing of free health care services by the patients. The study reports that majority of the population received diagnosis and treatment services for free of cost and that had significant impact on out-of-pocket expenditure. This also indicates awareness regarding available services and trust in the public health care system.

Though the study showed some of the important insights into the characteristic features of secondary infection of COVID-19 in household contacts in specific population of India, the generalizability of the findings needs to be validated in the different populations before recommending any policy decisions.

## Conclusion

Current study provided 1) SAR data from Gandhinagar rural district of Gujarat and compared it against global and national statistics. 2) critical information regarding greater susceptibility of the primary cases for poorer outcome as compared to secondary cases. 3) Policy implication that in an epidemic, government has provided top quality free services and hence no cost of diagnosis and treatment incurred to majority of the patients. That has showed drastic reduction in out-of-pocket expenditures and reduced cost burden on patients. However more robust studies with larger sample size are needed to substantiate findings of the current study and identifying epidemiological features of disease transmission.

## Data Availability

All the data is available with the authors and can be provided in case of peer-review.

